# Rett Syndrome in Ireland: A demographic study

**DOI:** 10.1101/2023.02.13.23285763

**Authors:** Komal Zade, Ciara Campbell, Snow Bach, Hazel Fernandes, Daniela Tropea

## Abstract

Rett syndrome (RTT) is a rare neuropsychiatric condition associated to mutations in the gene coding for the methyl-CpG-binding protein 2 (*MECP2*). It is primarily observed in girls and affects individuals globally. The understanding of the neurobiology of RTT and patient management has been improved by studies that describe the demographic and clinical presentation of patients with RTT. However, in Ireland, there is a scarcity of data regarding patients with RTT, which impedes the ability to fully characterize the Irish RTT population. Together with the Rett Syndrome Association of Ireland (RSAI), we prepared a questionnaire to determine the characteristics of RTT patients in Ireland. Twenty families have participated in the study to date, providing information about demographics, genetics, familial history, clinical features, and regression. The main finding of this study is the limited number of genetic tests conducted to support the clinical diagnosis of RTT. The results shows that Irish patients with RTT have comparable presentation with respect to patients in other countries, however, they had a better response to anti-epileptic drugs and fewer skeletal deformities were reported. Nonetheless, seizures, involuntary movements and regression were more frequently observed in Irish patients. Despite the limited sample size, this study is the first to characterise the RTT population in Ireland and highlights the importance of genetic testing for patients with RTT in order to sharpen the characterization of the phenotype and increase the visibility of Irish patients in the international RTT community.

## 1. Introduction

Rett Syndrome (RTT) is a rare neurodevelopmental disorder that affects brain function and development in approximately 7.1 per 100,000 females globally [1]. After an apparently typical development, patients with classic RTT present with regression of acquired developmental skills at 1-3 years of age. This period of regression is defined by the loss of babble and communicative behaviour, decrease of practical hand use and a decline in motor and intellectual ability of the individual [2].

RTT manifestations can range in clinical severity and are characterised by both mental and physical disabilities. There are four stages of RTT defining the clinical course: (i) Early onset: birth to 6-18 months, (ii) Rapid developmental deterioration: 1-4 years, (iii) Plateau/pseudo-stationary: 4-10 years, (iv) Late motor deterioration: 10-30 years [3].

The child with RTT exhibits apparently typical development until 6-18 months old, with some individuals experiencing developmental delays. Symptoms of Classic RTT include acquired hypotonia and microcephaly observed around 3-6 months of age. Loss of previously acquired skills and linguistic skills, poor muscle tone, slower cerebral and physical development, jerky limb movements, behavioural and emotional dysregulation like restricted sociability are observed around 1-4 years of age. Stereotypic hand movements are a defining feature of RTT. Other symptoms include seizures, bruxism, apraxia, cold hands and feet, gastrointestinal issues like bloating, cardiac and breathing issues (mainly episodic hyperventilation, and breath-holding, Valsalva manoeuvres); forced saliva and air expulsion, sleep disturbances, musculoskeletal abnormalities (predominantly scoliosis and lower limb deformities) [2, 4]. Autistic traits are present, but only in stage 2.

The majority of patients with RTT have mutations in the gene coding for Methyl-CpG binding protein 2 (*MECP2*) located on the Xq28 chromosome. RTT is X-linked, predominantly affecting females; males with RTT typically do not survive past a year [5]. MeCP2 is a chromosome-binding protein selective for 5-methyl cytosine residues generally located in the promoter region of genes that undergo transcriptional silencing following DNA methylation. Ultimately, this protein has a role in transcriptional repression via its transcriptional repression complex, and in transcriptional inhibition [6]. However, MeCP2 can also be an activator of gene transcription depending on the cofactors that are in the complex. *MECP2* is important for brain development and function, but the protein is ubiquitously expressed in the body. A study carried out in mice suggests that while RTT-associated symptoms mostly arise from a lack of *Mecp2* in the nervous system, some less extreme aspects of RTT can also occur independently of nervous system defects [7]. MeCP2’ s role has also been linked to the regulation of miRNA function [8]. The relationship between MeCP2 and miRNA, and its association with RTT is being investigated through studies on the impact of MeCP2 loss on miRNA expression and the potential involvement of MeCP2 domains and Rett-associated mutations in miRNA alterations [8]. Although over 90% of RTT patients have mutations in *MECP2* gene, there are some patients with a clinical diagnosis of RTT who do not have confirmed *MECP2* mutations. Recently, additional factors have been suggested to play a role in the development of RTT’s phenotype [9]. In addition to *MECP2*, other genetic variants have been identified as contributing to RTT-like phenotypes, including *CDKL5* and *FOXG1, MEF2C, TCF4* [10, 11], *CTNNB1, WDR45* [12], *ATP6V0A1, USP8, MAST3, NCOR2* [13].

RTT has several subtypes including classic/typical RTT, atypical RTT, and Rett-like. Atypical RTT has specific variants, such as the preserved speech or “Zappella variant” (Z-RTT), which is defined by milder symptoms and some speech ability. The classification of RTT and its subtypes is continually evolving, exhibiting a diverse clinical and genetic spectrum, and other syndromes were previously part of the RTT spectrum until recently: the “Hanefeld variant” (related to the *CDKL5* gene) is characterised by early onset seizures. The “Congenital variant” (related to the *FOXG1* gene) presents RTT symptoms from birth. *CDKL5* disorder and *FOXG1* disorder are now recognized as separate disorders with unique clinical and molecular characteristics [14, 15]. ‘Rett-like’ subtype refers to conditions similar to classic RTT, but with varying causes and genetic mutations, and not fulfilling all diagnostic criteria [16].

Overall, there is significant clinical and phenotypic variability associated with RTT. The variation in RTT presentation is associated with various factors, including: the nature of *MECP2* mutation present [17-20], the degree of X-chromosome inactivation [21, 22], the individual’s genetic background; and the site and location of MeCP2 expression in the individual’s brain [3]. Additional variability comes from the reports of demographic analysis from specific populations in several countries such as USA and Canada [23-28]; Italy [4, 29]; Denmark [30-32]; Australia [5, 33-39]; UK [36, 40-42]; the Netherlands and Belgium [43, 44]; Brazil [45]; Poland [46]; Sweden [47]; and an international analysis [48]. Due to this degree of variability in presentation, RTT is commonly misdiagnosed and often difficult to treat effectively, especially when considering differences in the genetic basis of the condition. The barriers resulting from the variability of RTT have led to the development of several RTT databases as a means to categorise and analyse different RTT cohorts. These include: InterRett, Australian Rett Syndrome Database (ARSD), Rett Syndrome Natural History Study (USA), and the Rett Networked Database [49].

These databases have led to great advancements in understanding the biology of RTT and in recording of genotypic and phenotypic variation of RTT. They also contributed to recognize the presence of different cohorts of patients. The primary advantage of recognizing a population cohort is to advance the criteria for patients’ selection in clinical studies. For example, in 2000 all patients enrolled in the ARSD were offered genetic testing for *MECP2* mutations, as the genetic mutation is an important parameter to be considered for patients’ stratification [33]. However, the genetic information is not always available for patients with RTT in Ireland. This missing information would be crucial for the proper diagnosis and characterization of the Irish RTT population and for advancing RTT research.

This is the first study investigating the Irish RTT population. This study analyses data from the Irish RTT population on the basis of information provided by the caregivers of patients with RTT. In collaboration with the Rett Syndrome Association of Ireland (RSAI), we prepared a set of questions aimed at capturing demographic information. The questionnaires were distributed, collected, and anonymised by the RSAI. The data was then analysed by researchers who were blinded to the participants’ identities. In the study the term “patient” refers to the individual diagnosed with Rett Syndrome; “participant” refers to the patient’s caregiver. The participants answered the questions referring to the patient in their care. Each participant corresponds to one patient only. We derived an overview of the general demographic of RTT patients in Ireland and compared it with data from RTT populations in other countries.

## 2. Materials and Methods

### 2.1 Study Design and Data Source

The primary objective of the RSAI is to enhance recognition and understanding of RTT in both clinical and public spheres, and to foster and support RTT research initiatives. Therefore, in collaboration with the association, we prepared a questionnaire for caregivers that outlines the demographic characteristics of RTT patients in Ireland. The questions are a subset of the questionnaire established by *Grillo et al*. and have been selected in partnership with the families affiliated to RSAI to create a user-friendly version that is comprehensible and easy for them to complete [49].

RSAI distributed questionnaires at the Belfast Regional Day event in May 2022. This event was a gathering for Irish and British patients with RTT and was jointly organised by Rett UK and RSAI. At the event, RSAI provided information on the purpose of the research and the benefits of participation and distributed the questionnaires, along with consent forms and patient information leaflets. In addition, the association disseminated information through a pre-recorded video and distributed the questionnaire to its remaining members. After collecting the questionnaires, RSAI removed personal information, assigned codes, and sent the anonymous data to the researchers for analysis. The entire process complied with General Data Protection Regulation policies and was approved by the TCD Faculty of Health Science Ethical Committee (210404). Only participants who provided consent were included in the analysis.

The questionnaire includes 26 questions, which inquired about the following details: Current age of patient; height of patient; weight of patient; age of diagnosis; place of diagnosis; clinician/service that performed the diagnosis; Type of RTT; genetic test and mutation; regression (behavioural, speech, motor, growth failure, and regression of hand use); hand use of patient; presence of gastrointestinal (GI) problems; communication (verbal and/or non-verbal); Motor skills of patient (ability to sit, walk, stand, walk with support) and ability to understand/interact with others; epilepsy status (seizures, age of seizure onset, current medications, resistance to anti-epileptic medications, family history of seizures, and details of the seizures); breathing abnormalities; bloating; cold extremities; sphincter control; skeletal abnormalities; involuntary movements; bruxism; age of pubertal onset (if begun); additional diagnostic tests performed (EEG, Cardiac activity, breathing activity, imaging); family history (consanguinity between the parents, history of RTT/other diseases in the family, siblings and their age, sex); mother’s pregnancy details (regular/irregular pregnancy, medications taken during the gestation period, regular/assisted delivery). For some questions, the participants were asked to choose the most commonly observed presentation from a list of options. For example, when asked about behaviour, the options included excitement, sadness, self-injury, injury to others, anxiety, and difficulty falling asleep at night. A blank space was left for patients to enter any additional notes which they felt were important to include in the questionnaire.

For the comparisons with other populations, the data was acquired from a literature search, using the search terms ‘Rett syndrome’, ‘RTT, phenotype’, ‘mutation’, ‘population’, and ‘international data’. Articles published in the years ranging from 1985 to 2022 were then chosen from a PubMed search. The nations in this comparison include the USA and Canada [23-28]; Italy [4, 29]; Denmark [30-32]; Australia [5, 33-39]; UK [36, 40-42]; the Netherlands and Belgium [43, 44]; Brazil [45]; Poland [46]; Sweden [47]; and an international analysis [48]. The countries are represented using the ISO country codes: USA-US, Italy-IT, Denmark-DK, Netherlands-NL, Sweden-SE, Australia-AU, UK-UK, Brazil-BR, Poland-PL, Ireland-IE; for the international cohort, we used abbreviation ‘Intl’.

### 2.2 Sample

The study sample includes only the participants who have answered the questionnaire to date, who were diagnosed with RTT and were recruited through RSAI. The analysis was conducted on all the questionnaires which were returned to the researchers at time of data analysis and submission (n=20). Additional questionnaires may be added in the future.

### 2.3 Organization and Analysis of the data provided by the Irish cohort

Data collected from the Irish Patient Cohort was cross-tabulated, based on the questions asked in the questionnaire. It was then transferred electronically into a database, then processed by software such as MS-Excel, Matplotlib 3.5.2, Python 3.8, NumPy 1.21.0, Seaborn, and Minitab 20. The numerical data was analysed by calculating the mean of all input provided (age, weight, height, etc). The non-numerical data was assessed by calculating the percentage of participants which answered ‘yes’ to the presence of the feature in the question.

### 2.4 Comparison between data from Irish RTT population and other cohorts from other countries

A binomial test was used to compare the categorical variables associated with the clinical signs and symptoms displayed by our sample, to populations in other countries. A two-sample t-test was carried to compare the numerical variables associated with the clinical signs and symptoms displayed by our sample, to populations in other countries. This analysis was carried out to assess if there was significant difference between the cohorts. For all analyses, significance was set at (p ≤ 0.05). The p-values adjustments were carried out implementing the Bonferroni correction method.

## 3. RESULTS

### Analysis of the Rett syndrome demographic data in Ireland

In collaboration with the Rett Syndrome Association of Ireland (RSAI), we prepared a questionnaire adapting some of the questions prepared by *Grillo* and colleagues that was distributed to the caregivers who were members of RSAI and who assisted a child with RTT [49]. The families were asked to complete the questionnaire, participant information leaflet and consent form jointly. The families returned these documents to the RSAI who anonymised them, removed personal contacts, and forwarded these to the researchers for the analysis. At the time of this submission twenty questionnaires were received and used for analysis. They are indicative of the trends in symptom presentation seen in the Irish RTT population. To facilitate the analysis, we organised the questionnaire in different sections: General information; Diagnosis; Regression of acquired skills; Presentation of symptoms-gastrointestinal probIems, communication, motor skills, epilepsy, breathing abnormalities, cold extremities, skeletal abnormalities, involuntary movements, bruxism; Behaviour; Additional diagnostic testing; Family details of RTT patients in Ireland; and pregnancy details.

#### 3.1 General Information

In questions 1-4, we collected general information that is summarised in *Table 1*. The participants report that the age of diagnosis was 5.21 ± 7.043 years and the pubertal onset was observed at the age of 10.81 ± 1.703 years. Some participants report using medication to delay puberty, while others report that the patients have not yet started menstruating despite experiencing the onset of puberty. Most of the patients in the study are young adult females, and their height, weight and pubertal onset age are proportional to their age.

**Table 1.** General Information on the patients with RTT in Ireland. The tabular data shows the participants’ demographics, including mean and standard deviation for their age, height, and weight at the time of the study, the age at diagnosis and age of pubertal onset. For the analysis, we only consider the answered responses as some questions were left unanswered.

| **Demographics** | **Mean ± SD** | **Range** |
| --- | --- | --- |
| Current age | $20.6 \pm 11.748$ years | 2 to 46 years |
| Current height | $4.9 \pm 0.468$ feet | 4 to 5.7 feet |
| Current weight | $37.67 \pm 14.354$ kg | 10 to 65 kgs |
| Age of diagnosis | $5.21 \pm 7.043$ years | 1.1 to 30 years |
| Age of puberty onset | $10.81 \pm 1.703$ years | 8 to 14 years |

#### 3.2 Diagnosis

Questions 5-8 focused on the diagnostic information reported as classic or atypical RTT diagnosis reported in *Figure 1*. The study sample consists of a mix of patients with reported typical and atypical cases, with some cases lacking a diagnosis.

**Figure 1.**
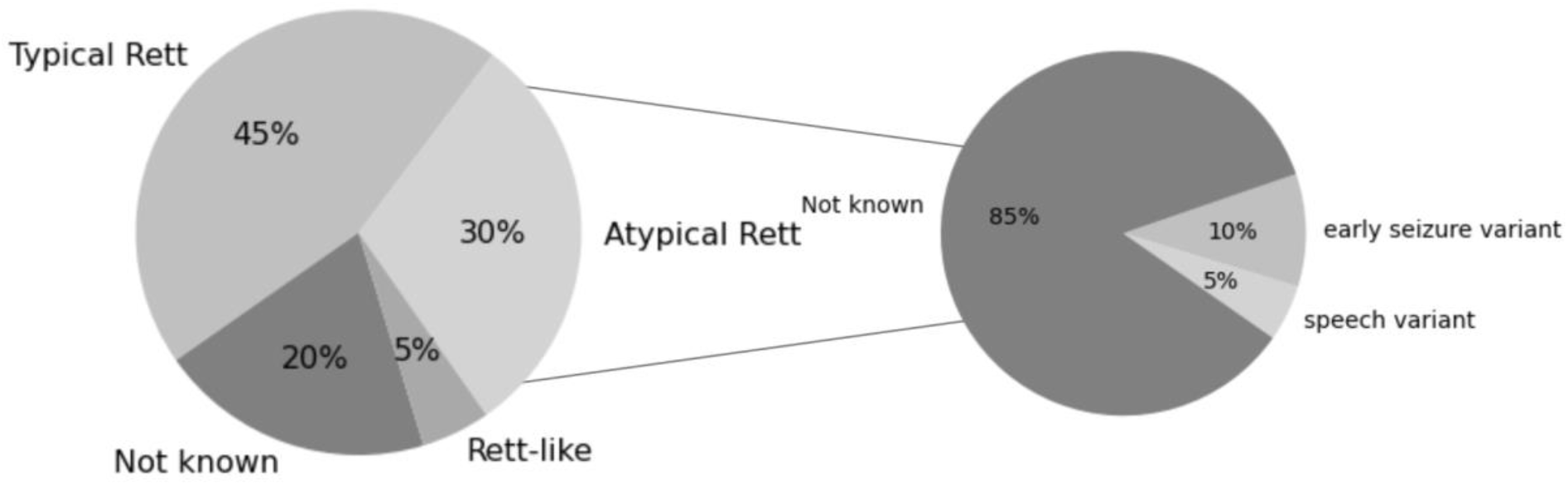
Reported information on the different clinical RTT form RTT diagnosed in patients in Ireland. The larger pie chart shows the percentage of different clinical RTT forms-typical, atypical and Rett-like as reported in our study cohort. The smaller pie chart shows the different forms of atypical RTT reported by the participants.

For the information on type of Rett, 45% (n=9) reported to have a diagnosis of typical RTT and 30% (n=6) reported to have a diagnosis of atypical RTT. 5% (n=1) participants reported their RTT subtype as ‘Rett-like’ and 20% (n=4) did not answer this question. Among the families that reported a diagnosis of atypical RTT, 5% (n=1) reported to have speech variant, and 10% (n=2 reported to have ‘the early seizure variant’. Genetic test was performed in 85% (n=17) of patients, and not performed in 10% (n=2), while 5% (n=1) did not answer. Despite the majority of the participants reported that the genetic test was performed, only 30% (n=6) of them detailed the mutation observed, 10% (n=2) reported general *MECP2* mutation without providing details. 60% (n=12) did not report any information regarding the mutation. The *MECP2* mutations described by the participants were: R306C, R133C, P152R, c.455C>G (p.Pro152Arg), deletion in exon 4, and deletion in exons 3 and 4.

Some participants also reported information related to the residence of the patients. All the patients were living at their home with family, except one individual (amongst the older participants) who was reported to live in a residential care facility. Participants provided information about the facilities/hospitals where they received their diagnosis. The frequency of diagnosis at specific hospitals are listed in *Figure 2*. The hospitals mentioned were Crumlin hospital, Cavan hospital, Temple Street, RVH Belfast, Belfast City Hospital (BUH), CUH Cork, Glasgow, Lucan, Tallaght hospital. Most patients had their clinical diagnosis made in Ireland, although their genetic diagnosis was obtained from the UK.

**Figure 2.**
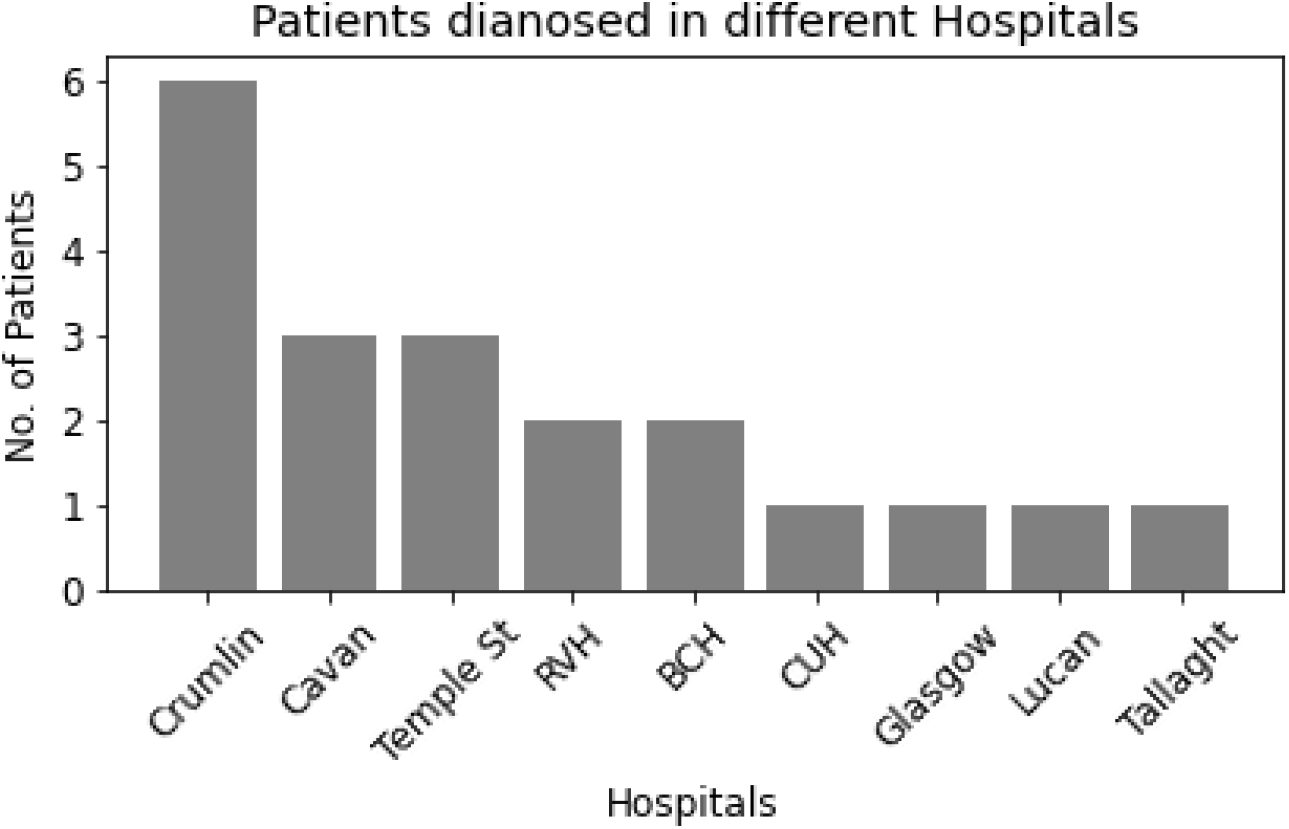
Hospital-wise diagnosis of the patients with RTT in Ireland. The bar graph represents the number of patients diagnosed at the facilities/hospitals in Ireland.

#### 3.3 Regression of acquired skills

Regression (Question 9), a common feature of RTT presentation, was reported based on the six parameters reported in *Table 2*. For general regression of acquired skills, 95% (n=19) participants reported ‘yes’, 5% (n=1) participants reported ‘no’. One of the participants reported no observed regression in the patient, which was the youngest in the cohort, and therefore regression might yet to be observed. 80% (n=16) participants reported that the regression started at the age of 1.54 ± 0.609 years, 20% (n=4) participants did not answer to this question. For behavioural regression, 65% (n=13) participants reported ‘yes’, 35% (n=7) participants reported ‘no’. 60% (n=12) participants reported that the behavioural regression started at the age of 1.54 ± 0.609 years, 40% (n=8) participants did not answer to this question. Regarding speech regression, only 5% (n=1) of cases reported ability to speak using a few words, the remaining 95% (n=19) patients reported inability to speak. In relation to regression of hand use, 85% (n=17) reported ‘yes’, 10% (n=2) reported ‘no’, and 5% (n=1) did not answer. The participants provided information about the patient’s current hand use, 90% (n=18) reported lost normal hand use and 5% (n=1) reported normal hand use and 5% (n=1) did not answer. 30% (n=6) reported poor grasp, 10% (n=2) reported one hand to be more functional than the other. A substantial number of participants reported regression in speech, motor skills, and hand use, while regression in behaviour and growth was less prevalent.

**Table 2.**
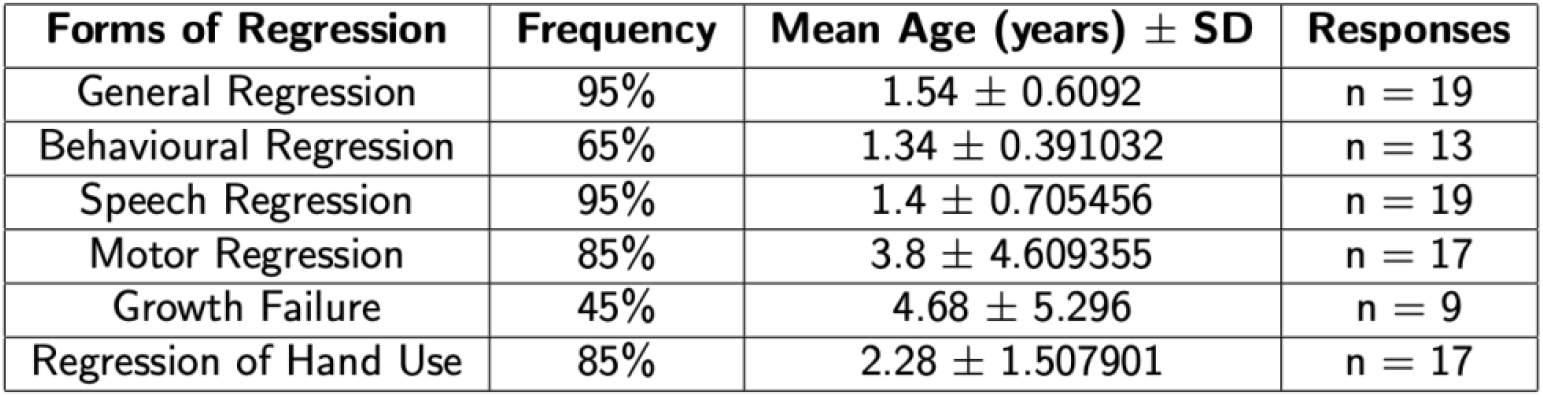
Regression with their forms observed in the patients with RTT in Ireland. The tabular data shows the frequency of regression reported by the participants including general regression, regression in behaviour, speech, motor, growth failure, and hand use. Age (average and standard deviation) at which specific forms of regression were observed, the number of responses received for each parameter are mentioned.

| Forms of Regression | Frequency | Mean Age (years) ± SD | Responses |
| --- | --- | --- | --- |
| General Regression | 95% | $1.54 \pm 0.6092$ | n = 19 |
| Behavioural Regression | 65% | $1.34 \pm 0.391032$ | n = 13 |
| Speech Regression | 95% | $1.4 \pm 0.705456$ | n = 19 |
| Motor Regression | 85% | $3.8 \pm 4.609355$ | n = 17 |
| Growth Failure | 45% | $4.68 \pm 5.296$ | n = 9 |
| Regression of Hand Use | 85% | $2.28 \pm 1.507901$ | n = 17 |

#### 3.4 Presentation of symptoms

Questions 10-22 inquired about specific symptoms: Gastrointestinal problems, communication, motor skills, epilepsy and other clinical presentations, the results are indicated in *Table 3*.

**Table 3.** Details of symptomatic presentation of the patients with RTT in Ireland. The tabular data shows the frequency of issues like gastrointestinal, motor, additional clinical presentation, communication, epilepsy; the number of responses received for each parameter are mentioned.

| Problems | Frequency (%) | Responses |
| --- | --- | --- |
| **Gastrointestinal issues** |  |  |
| General gastrointestinal problems | 90 | n = 18 |
| Constipation | 75 | n = 15 |
| Reflux | 35 | n = 7 |
| Chewing and swallowing problems | 40 | n = 8 |
| Peg fed | 15 | n = 3 |
| Bloating | 60 | n = 12 |
| Sphincter Control | 35 | n = 7 |
| **Communication** |  |  |
| Verbal Communication | 10 | n = 2 |
| Non-Verbal Communication | 75 | n = 15 |
| **Motor skills** |  |  |
| Ability to sit | 50 | n = 10 |
| Ability to walk | 20 | n = 4 |
| Ability to stand | 55 | n = 11 |
| Walking with support | 50 | n = 10 |
| Ability to understand and interact with others | 65 | n = 13 |
| **Epilepsy** |  |  |
| Current Medications | 85 | n = 17 |
| Seizures | 85 | n = 17 |
| Resistance to Medications | 15 | n = 3 |
| Family History of Seizures | 15 | n = 3 |
| **Other Clinical Presentations** |  |  |
| Breathing abnormalities | 65 | n = 13 |
| Cold Extremities | 80 | n = 16 |
| Skeletal abnormalities | 55 | n = 11 |
| Involuntary movements | 85 | n = 17 |
| Bruxism | 50 | n = 10 |

##### Gastrointestinal problems

Gastrointestinal issues (Question 11) were reported by 90% (n=18) of the participants and 10% (n=2) did not report them. Regarding the specific gastrointestinal problems, constipation was reported in 75% (n=15) and bloating in 60% (n=12) of the patients. 35% (n=7) of the patients could control their sphincter muscle, while 40% (n=8) could not, and 25% (n=5) of the participants did not answer. Reflux was reported in 35% (n=7) of the patients, chewing and swallowing difficulties in 40% (n=8), and 15% (n=3) of the patients were fed through a feeding tube known as a PEG-percutaneous endoscopic gastrostomy (*Table 3*). Constipation and bloating were the most prevalent gastrointestinal problems reported by the participants, while other issues such as reflux, chewing difficulties, and swallowing problems were less common.

##### Communication

For communication (Question 12), 75% (n=15) of participants reported patients to engage in non-verbal communication and 10% (n=2) reported patients to engage in verbal communication, and 15% (n=3) reported no communication *(Table 3)*. The reported non-verbal communication behaviours were 65% (n=13) Communication with eye contact, and 25% (n=5) communication through either babble speech, mobilisation, shouting for attention, or turning their head to indicate ‘no’. 5% (n=1) patients maintained full speech and were capable of verbal communication while another 5% (n=1) had limited speech. The individual who fully maintained speech was reported to have the preserved speech variant, while the individual with limited speech was reported to have the early seizure variant. Most of the participants reported that the patients primarily used nonverbal communication, with eye contact or gazing being the most common form.

##### Motor skills

The motor skills of patients (Question 13) were assessed based on their abilities to sit, stand, walk with support, walk without support, to understand and interact with others. The resulting data from these questions are represented in *Table 3*. The results from this section indicate a comparatively high percentage of the patients in the study 65% (n=13) maintained the ability to understand and interact with others, despite the lack or decreased functional skills. It was unclear in 5% (n=1) whether they possessed the ability to understand and interact with others. 10% (n=2) participants reported the patient’s ability to stand with support, and 5% (n=1) needed support with walking during periods of regression. 5% (n=1) patient’s ability to walk/stand was lost at ∼5 years old. A substantial number of participants were able to understand and interact with others, and about half of them could stand, sit, and walk with assistance. Only 20% (n=4) were able to walk independently.

##### Epilepsy

The epileptic status (Question 14) was enquired in relation to the parameters exhibited in *Table 3*. 85% (n=17) of the participants reported that the patients experience epileptic seizures from the age of 7.39±4.657 years. Seizures were described in detail only by 60% (n=12) of participants, who reported that 20% (n=4) of patients exhibited tonic clonic seizures, and 40% (n=8) exhibited focal, absence or partial seizures (*Figure 3)*. For one patient the epilepsy onset was associated with the onset of menstruation. The participants reported that 85% (n=17) of the patients were taking medications for epilepsy and 15% (n=3) did not respond to anti-epileptic drugs. A family history of temporal lobe epilepsy was reported for 5% (n=1), an unspecified family history of epilepsy was reported for 10% (n=2) of the patients. In summary, the prevalence of seizures was high among the patients, with few of them being resistant to medications.

**Figure 3.**
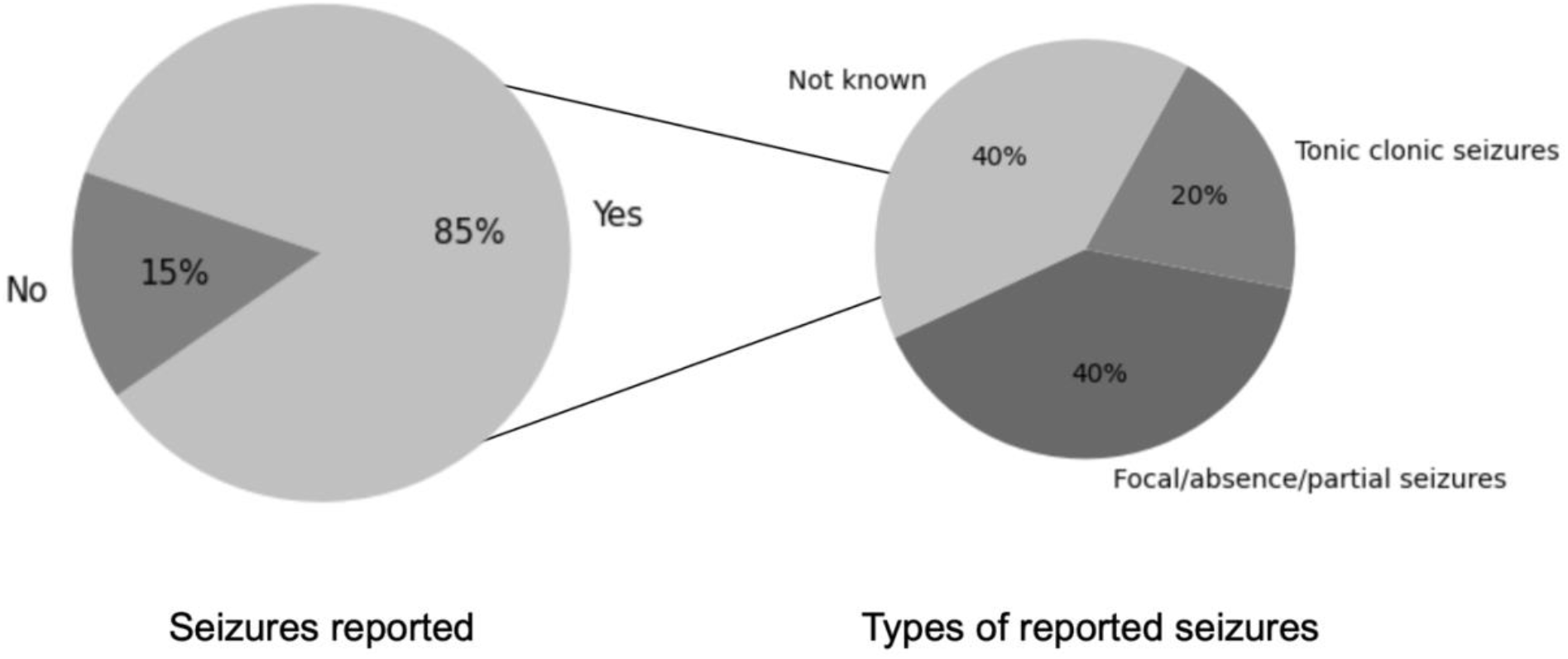
Reported information on the frequency of seizures and forms of seizures reported in patients in Ireland. The larger pie chart shows the percentage of seizures as reported in our study cohort. The smaller pie chart shows the different forms of seizures reported by the participants.

##### Breathing abnormalities

Participants reported breathing abnormalities (Question 15) in 65% (n=13) of the patients. They included episodes of rapid breathing, irregular breathing patterns, and apnoea.

##### Cold Extremities

One of the common symptoms of RTT is cold extremities, reported by 80% (n=16) of the participants. The decreased blood flow to the hands and feet led to decreased dexterity and muscle tone in the hands.

##### Skeletal abnormalities

55% (n=11) of the patients were reported to have skeletal abnormalities (Question 19) including scoliosis. These abnormalities caused pain and discomfort and affected the patient’s ability to walk and move. There were no skeletal abnormalities in 40% (n= 8) of the patients. 5% (n=1) of the participants did not respond to this question. A patient underwent scoliosis surgery.

##### Involuntary movements

Involuntary movements (Question 20) were reported in in 85% (n=17) of the patients; they included hand wringing, repetitive finger movements, and irregular movements of the limbs and trunk. There were no reports of involuntary movement in 10% (n=2) of the patients, and 5% (n=1) of the participants did not answer the question.

##### Bruxism

Bruxism (Question 21), another feature of RTT, was reported in 50% (n=10) of the patients while 30% (n=6) did not present bruxism and 20% (n=4) of the participants did not answer the question.

To summarise, stereotypic involuntary movements and cold extremities were commonly reported by the majority of participants, while breathing issues, skeletal problems, and bruxism were found to be less prevalent, as summarised in *Table 3*. In addition to these symptoms, eosinophilic oesophagitis was reported in one patient.

#### 3.5 Behaviour

The behaviours most often observed in our patient cohort are represented in *Figure 4*. The most commonly reported behaviour observed (Question 23) was excitement in 85% (n=17), sadness in 80% (n=16), anxiety in 75% (n=15), night-time sleep disturbance in 55% (n=11), self-injury in 20% (n=4) and injury to others in 5% (n=1).

**Figure 4.**
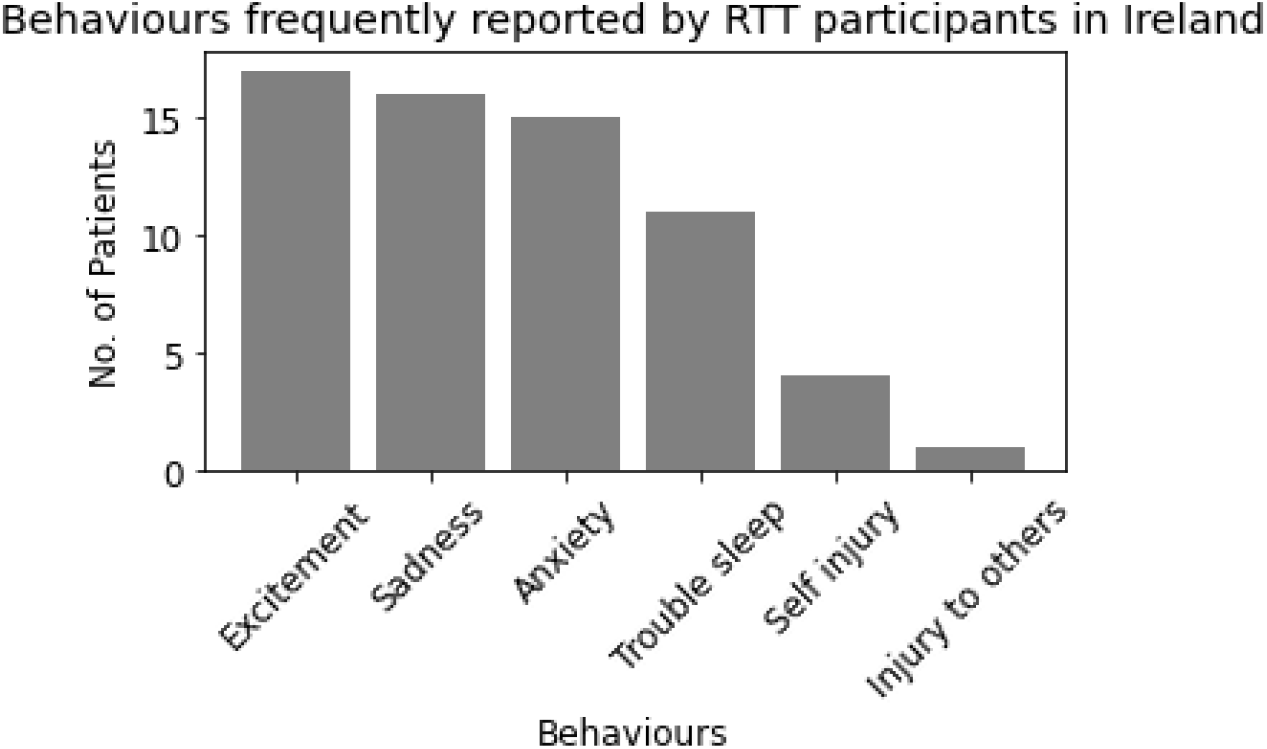
Details of behavioural presentation of the patients with RTT in Ireland. The bar chart shows the number of patients displaying the behaviours, such as excitement, sadness, anxiety, difficulty sleeping during night, self-harm, and injury to others.

#### 3.6 Additional diagnostic testing

The frequency of participants from our study who underwent additional diagnostic tests (Question 24) are represented in *Figure 5*. 85% (n=17) of the participants reported that the patients had an EEG test, 50% (n=10) had measurement of their breathing activity, and 45% (n=9) had evaluation of cardiac activity and imaging. Among the diagnostic tests conducted on the participants, EEG was the most widely performed test, while tests related to breathing activity, cardiac activity, and imaging were performed less frequently.

**Figure 5.**
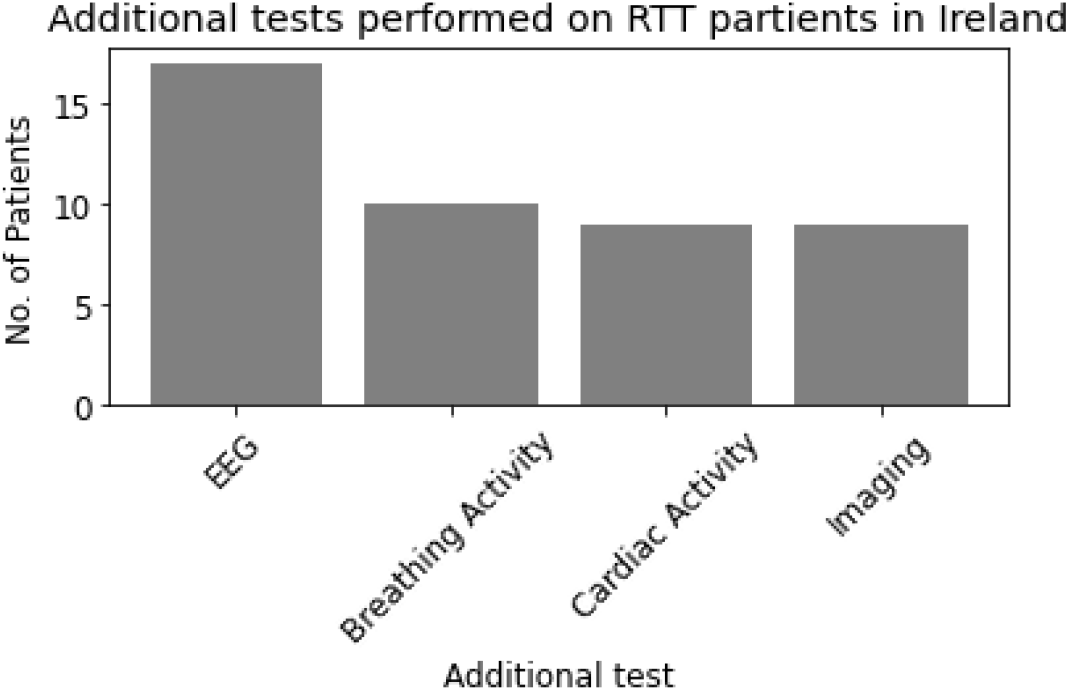
Details of diagnostic tests performed on the patients with RTT in Ireland. The bar chart shows the number of patients who underwent the additional diagnostic tests including EEG, Cardiac activity, Breathing activity and Imaging.

#### 3.7 Family details of RTT patients in Ireland

The participants were asked about family data (Question 25) such as the presence of consanguinity (between parents); a family history of RTT; other diseases present in the family; and number, age, and sex of siblings - if present. A family history of other diseases including Heart Disease, Thyroid Disease, Depression, Psoriasis, Psoriatic Arthritis, Multiple Sclerosis, Autism, Anxiety, and Type II Diabetes was reported by 35%(n=7) of the participants. 85% (n=17) of participants reported that the patients had siblings; of those 75% (n=15) shared information about the sex of the patient’s siblings. Mean age of the patient’s siblings was 21.41 years. There was no family history of RTT reported. Consanguinity between parents of the patient was reported by 5% (n=1) of the participants. Most of the patients had siblings and approximately half of participants reported a family history of various other ailments but not RTT.

#### 3.8 Details of pregnancy

5% (n=1) of the participants reported multiple pregnancy for the details of pregnancy (Question 26). 10% (n=2) of the mothers had taken medications, including carbamazepine and heparin during the pregnancy. Assisted delivery interventions were reported in 45% (n=9) of the cases, and Caesarean deliveries accounted for 7 of the 9 assisted delivery interventions. Other complications reported were the use of forceps, face presentation, intrauterine and post-birth blood transfusion, and obstructed labour. Nearly half of participants had assisted deliveries, the majority of which involved C-sections.

### Analysis of the population data of patients with RTT in Ireland and comparison with other countries

#### 3.9 Representation of parameters in populations with RTT across several countries

To compare the data from Ireland to those of other countries, we considered variables such as current age and weight of the patient at the time of the study, the age of diagnosis, the age at which regression was observed and the age of seizure onset. *Table 4* shows the data in each population. The literature available so far does not include data for the listed variables across all the populations.

**Table 4.** Representation of the data in the RTT cohort of Ireland and to other countries. The tabular representation of data such as the current age and weight of the patients, age of diagnosis, age of regression observed, and age of onset of seizures in the study cohort in Ireland and other international cohorts, such as the USA, Italy, Denmark, Netherlands, Australia, UK, Brazil, and Poland. The numbers in the figure are accompanied by asterisks to indicate the level of significance of the differences from Ireland (IE). A single asterisk (*) indicates a significant difference from Ireland for the indicated distinctive parameter. The double asterisks (**) indicate a higher level of significance, determined following p-value adjustments for multiple testing. The numbers shown in bold for Ireland indicate an extreme end parameter that has been reported in Ireland and differs greatly from other countries.

| Characteristic | Mean Value within Sample |  |  |  |  |  |  |  |  |  |  |
| --- | --- | --- | --- | --- | --- | --- | --- | --- | --- | --- | --- |
|  | US | IT | DK | NL | SE | AU | UK | BR | PL | INTL | IE |
| Age (years) | 3** | 29** | 40.7** | 14.9* | 19.6 | 13.7** | 20.5 | 12.2** | 7.22** | - | 20.6 |
| Current Weight (kg) | - | - | 42 | - | - | - | - | - | - | - | 37.7 |
| Age of Diagnosis (years) | 2** | - | 2** | 7.8** | - | 5.5 | 3** | - | 3.5 | - | 5.2 |
| Age of Regression (years) | - | - | - | 1** | - | 2.6** | 1.5 | - | - | - | 1.5 |
| Age of Seizure onset (years) | - | 5* | - | 4 | - | 3.4** | - | - | - | - | 7.4 |

To compare the RTT characteristics data between Ireland and other populations, we considered the frequency of parameters including the presence of general regression, behavioural regression, speech, motor skills, hand use, stereotypies, gastrointestinal issues, communication including verbal and nonverbal, eye communication, the patient’s ability to sit, stand and walk (with and without support), epilepsy (seizures), resistance to anti-epileptic drugs, breathing abnormalities, bloating, cold extremities, sphincter control, skeletal abnormalities, involuntary movements, bruxism, behaviours like anxiety, self-injury, trouble night sleep, and sadness. *Table 5* represents the data in each population.

**Table 5:** Frequency of RTT characteristics in the Irish population compared to patients in other countries. The tabular representation of RTT characteristics in the study cohort in Ireland and other international cohorts, from the USA, Italy, Denmark, the Netherlands, Sweden, Australia, the UK, Brazil, Poland, and an international cohort. The numbers in the figure are accompanied by asterisks to indicate the level of significance of the differences from Ireland (IE). A single asterisk (*) indicates a significant difference from Ireland for the indicated distinctive symptom. The double asterisks (**) indicate a higher level of significance, determined following p-value adjustment for multiple testing. The numbers shown in bold for Ireland indicate an extreme end frequency that has been reported in Ireland and differs greatly from other countries. This table provides a visual representation of the differences in the frequency of occurrence of distinctive symptoms between the cohort and Ireland and highlights the significant differences and extreme end frequencies.

| Characteristic | Percentage of Sample with Characteristic |  |  |  |  |  |  |  |  |  |  |
| --- | --- | --- | --- | --- | --- | --- | --- | --- | --- | --- | --- |
|  | US | IT | DK | NL | SE | AU | UK | BR | PL | INTL | IE |
| Regression | 90 | - | - | - | - | 76 | 95.6 | - | - | 80 | 95 |
| Behavioural Regression | - | - | - | - | - | - | 77 | - | - | - | 65 |
| Speech Regression | 87 | 50 | - | - | 23 | - | - | - | - | - | 95 |
| Motor Regression | 55 | - | - | - | - | - | - | - | - | - | 85 |
| Regression of Hand Use | - | - | 41 | 41 | - | - | - | - | - | - | 85 |
| Stereotypies | 86 | 98 | - | 96 | - | 92 | 99* | - | 70 | 69 | 85 |
| GI problems | 92 | 89 | - | - | - | 83 | 82 | 81 | - | 40 | 90 |
| Verbal Communication | - | 16 | - | 13 | - | 5 | 21 | - | - | - | 10 |
| Non-verbal Communication | - | 76 | - | 86 | 65 | 39 | 79 | - | - | 70 | 75 |
| Eye Communication | - | - | 85 | - | - | 92 | 91 | - | - | 89 | 65 |
| Ability to Sit | - | 37 | 85** | - | - | - | - | 59 | - | - | 50 |
| Ability to Walk | - | 20 | 44 | 33 | 80 | 56 | 66 | 22 | - | 44 | 20 |
| Ability to Stand | - | - | - | - | - | 46* | 28 | - | - | - | 55 |
| Walking with Support | - | 41 | 41 | - | - | 43 | 64 | 11 | - | - | 50 |
| Seizures | 81 | 82 | 80 | 61 | - | 81 | 63 | 69 | 43 | 65 | 85 |
| Resistance to Anti-epileptics | - | 36 | 46* | 18 | - | 36 | - | - | - | 25 | 15 |
| Breathing Abnormalities | - | 70 | 75 | 81 | 85* | - | 66 | - | 87 | 51 | 65 |
| Bloating | - | - | - | - | - | 53 | - | - | - | - | 60 |
| Cold Extremities | - | 87 | - | 56 | 93* | - | - | - | - | 50 | 80 |
| Sphincter Control | - | - | - | - | - | - | 30 | - | - | 89 | 35 |
| Skeletal Abnormalities | 27 | 56 | 83* | 70 | 74 | 66 | 67 | 59 | 13 | 50 | 55 |
| Involuntary Movements | - | - | - | 40 | - | - | - | - | - | - | 85 |
| Bruxism | - | - | - | - | - | - | - | - | 22 | 61 | 50 |
| Anxiety | 14 | - | - | - | - | 79 | 74 | - | - | - | 75 |
| Self-Injury | - | - | - | - | - | 62** | 57** | - | - | - | 20 |
| Trouble night sleep | - | 77 | - | 16 | - | 63 | 46 | - | - | 47 | 55 |
| Sadness | - | - | - | 12 | - | - | - | - | - | - | 80 |

#### 3.10 Frequency of different RTT forms in different populations

The distribution of different RTT forms in several population datasets, including those from the international cohort, United States, Italy, the Netherlands, Australia, the United Kingdom, Brazil, and Ireland, is shown in *Table 6*. The prevalence of RTT, which ranges between 68-85% for classical RTT and 13-35% for atypical RTT, is not consistent in Ireland between the classical and atypical forms.

**Table 6:** Distribution of RTT forms in Ireland compared to patients in the other countries. The tabular representation of RTT forms-classic/typical and atypical RTT in the study cohort in Ireland and other international cohorts, from the USA, Italy, Denmark, the Netherlands, Sweden, Australia, the UK, Brazil, Poland, and an international cohort. The figures in Ireland could be affected by the lack of diagnosis and genetic information.

| RTT Type | Percentage of sample with RTT Type |  |  |  |  |  |  |  |  |  |  |
| --- | --- | --- | --- | --- | --- | --- | --- | --- | --- | --- | --- |
|  | US | IT | DK | NL | SE | AU | UK | BR | PL | INTL | IE |
| Typical RTT | 85 | 65 | - | 80 | - | 68 | 76 | 68 | - | 80 | 45 |
| Atypical RTT | 13 | 35 | - | 20 | - | 32 | 21 | 27 | - | 20 | 30 |

## 4. DISCUSSION

This is the first study that provides information about demographic data among patients diagnosed with RTT in Ireland. In collaboration with the RSAI, we presented a questionnaire to the caregivers to gather information about clinical and behavioural presentation of Irish patients with RTT. The results indicate that patients in Ireland display better overall presentation, including less frequent skeletal deformities and greater motor abilities-although this last result could be due to the high number of patients with speech variant in our sample. The majority of the patients exhibit regression, seizures, and involuntary movements. The study reveals absence of genetic diagnosis in a significant number of patients, due to limited access to genotyping facilities and diagnostic services available to the public. This may lead to misdiagnosis of atypical RTT cases, as some participants report “early seizure variant of RTT” without being able to report the genetic mutation. It is possible that these patients may actually have ‘Hanefeld syndrome’, caused by *CDKL5* mutations, presenting similar symptoms but with early onset and frequent seizures [50].

We acknowledge that the sample size may not be representative of the entire Irish RTT population also considering that, according to the global prevalence of RTT (7.1 per 100000 females), we should expect about 180 patients with RTT in Ireland, while the RSAI only counts 75 members. Nonetheless, the results provide an insight into the phenotypic presentation of RTT patients in Ireland. The present study shows that clinical severity worsens with age, and there is a correlation between the delayed onset of RTT and the preserved hand function. This study includes both typical and atypical cases of RTT, with 30% of the participants reporting the atypical variant (but the percentage could be affected by the lack of proper genetic information). A small percentage of participants show excess weight for age and height, similar to children with Autism Spectrum Disorder [51]. The average age of RTT diagnosis in Ireland is 5.21 years, which is higher than in Poland, the UK, the US, and Denmark, but lower than in Australia and the Netherlands. Despite limited access to RTT diagnosis in Ireland, the average age of diagnosis seems to be comparable to that of other populations [5, 23].

RTT affects growth and is associated with issues like feeding difficulties, motor function, digestion, apraxia, hyperventilation, sleep, and scoliosis [52]. The results show a milder phenotype of scoliosis in patients who can walk unassisted and a trend of scoliosis not progressing in the p.Arg306Cys mutant patient. We also find that the behaviour worsens with age and that the younger patients display better mood, in line with previous literature [53]. Constipation is a majorly reported gastrointestinal issue. Some patients are PEG-fed or receive gastrostomy feed to meet their nutritional needs, which helps maintain their BMI [54]. While some of the parameters are comparable to other cohorts, RTT patients in Ireland display late onset of seizures; marked regression and involuntary movements. Although the majority of the participants report the patients taking medications for epilepsy, only a handful of them are resistant to anti-epileptics. Correcting Vitamin D deficiency can lower anti-epileptic drug dosage and improve health [55, 56]. Studies report low levels of Vitamin D in RTT patients were more prominent in summer than winter and can be related to less sun exposure from being indoors [56]. This literature confirms our finding that cold extremities are more prevalent in Sweden than in Ireland due to the country’s lower temperature [56]. Skeletal abnormalities and bone density can depend on Vitamin D deficiencies. Considered the number of patients with RTT presenting this problem it would be recommended to monitor bone density periodically. None of the patients in our study has a history of RTT and consanguinity between parents was reported only for one patient. 50% of participants report that the patients were delivered via C-sections, which has been reported to increase the risk of neuropsychiatric conditions [57].

We plan to increase the number of participants in the study in order to better characterise the RTT population in Ireland. The literature suggests that Irish individuals with RTT are currently not included in international studies or clinical trials. To address this, we are developing a database called ‘DATARETT’ that contains information about RTT patients in Ireland. The data obtained from our study will be input into DATARETT in compliance with GDPR regulations and ethics. The goal is to increase the representation of RTT in Ireland on a global scale, and to encourage future research and collaboration. Multiple existing centralised RTT databases have contributed to improving diagnosis, treatment, and future developments for RTT patients. The inclusion of Irish patients in such databases would be beneficial for RTT patients, scientists, healthcare professionals and the Irish community.

## Data Availability

The analysed data produced in the present work are contained in the manuscript while some supplemental data are available upon reasonable request to the authors.

## ACKNOWLEDGEMENT

The authors express their gratitude to the Rett Syndrome Association of Ireland (RSAI) for connecting them with patients and for their support throughout the study, and to all the participating families. We would like to thank the IRC New Foundation for their support and funding of this research project [IRCNF2021]. DT has been partially supported by IRSF (3507-207417 grant), Meath Foundation (Research award 2019), Fondation Jérôme Lejeune (DT Project#1935), and Science Foundation Ireland (SFI) under Grant Number 16/RC/3948 and co-funded under the European Regional Development Fund and by FutureNeuro industry partners.

